# Extended compartmental model for modeling COVID-19 epidemic in Slovenia

**DOI:** 10.1101/2022.07.16.22277702

**Authors:** Miha Fošnarič, Tina Kamenšek, Jerneja Žganec Gros, Janez Žibert

**Affiliations:** Faculty of Health Sciences, University of Ljubljana, Zdravstvena pot 5, SI-1000 Ljubljana, Slovenia; Alpineon, d.o.o., Ulica Iga Grudna 15, SI-1000 Ljubljana, Slovenia

**Keywords:** epidemiological modeling, Covid-19 epidemic in Slovenia, extended SEIR model, short-term forecasts, European Covid-19 Forecast Hub

## Abstract

In the absence of a systematic approach to epidemiological modeling in Slovenia, various isolated mathematical epidemiological models emerged shortly after the outbreak of the COVID-19 epidemic. We present an epidemiological model adapted to the COVID-19 situation in Slovenia. The standard SEIR model was extended to distinguish between age groups, symptomatic or asymptomatic disease progression, and vaccinated or unvaccinated populations. Evaluation of the model forecasts for 2021 showed the expected behavior of epidemiological modeling: our model adequately predicts the situation up to 4 weeks in advance; the changes in epidemiologic dynamics due to the emergence of a new viral variant in the population or the introduction of new interventions cannot be predicted by the model, but when the new situation is incorporated into the model, the forecasts are again reliable. Comparison with ensemble forecasts for 2022 within the European Covid-19 Forecast Hub showed better performance of our model, which can be explained by a model architecture better adapted to the situation in Slovenia, in particular a refined structure for vaccination, and better parameter tuning enabled by the more comprehensive data for Slovenia. Our model proved to be flexible, agile, and, despite the limitations of its compartmental structure, heterogeneous enough to provide reasonable and prompt short-term forecasts and possible scenarios for various public health strategies. The model has been fully operational on a daily basis since April 2020, served as one of the models for decision-making during the COVID-19 epidemic in Slovenia, and is part of the European Covid-19 Forecast Hub.

## 1 Introduction

The COVID-19 pandemic forced nations worldwide to suspend significant parts of their social and economic activities [1]. In Slovenia, SARS-CoV-2 was first detected in March 2020, and containment measures soon followed [2]. Especially before the development and rollout of vaccines against SARS-CoV-2, such non-pharmaceutical interventions were the only means available to countries to slow down coronavirus infection rates and avoid overburdening health care systems. Because of potentially high social and economic costs of containment measures, it is important to make informed decisions about when to implement them and at what scale. A useful tool for predicting and controlling the evolution of infectious diseases and understanding the impact of public health interventions is mathematical epidemiological modeling [3].

In the absence of a systematic approach to mathematical epidemiological modeling in Slovenia, various isolated mathematical epidemiological models emerged shortly after the outbreak of the COVID-19 epidemic. To benefit from more systematic access to epidemiological data and peer-review process, some of them [4–7] gathered around the emerging web portal COVID-19 Sledilnik [2]. Soon COVID-19 Sledilnik was widely accepted as a portal for aggregation, analysis, and representation of COVID-19 epidemiological data in Slovenia, and the results of some models [4,7] were used by government decision-makers.

In this work, we present an epidemiological model adapted to the COVID-19 epidemic in Slovenia [7]. The standard SEIR model [3] was extended to distinguish between age groups, symptomatic or asymptomatic disease progression, and vaccinated or unvaccinated populations. Similar extensions of SEIR-like models have been widely used in the COVID-19 crisis, for example, to account for undetected infections, different stages of infection or age groups [8–12], the effects of vaccination and coexistence of different viral variants [13], to study different behavioral responses to public health interventions [14,15], or to forecast burden of epidemics on health care systems [16].

The model presented in this paper has been fully operational on a daily basis since April 2020, served as a decision support tool during the COVID-19 epidemic in Slovenia, was used for COVID-19 Sledilnik data analyses [2], and has been included in the European Covid-19 Forecast Hub [17] coordinated by the European Centre for Disease Prevention and Control.

The rest of the paper is organized as follows. In the Methods section, the structure of the model with all its extensions, the outputs of the model, and the model evaluation methodology are explained. The model forecasts were evaluated for the years 2021 and 2022. The epidemic situation in Slovenia in 2021 and 2022 is described together with the evaluation results in the Results section and discussed in the following section. Before the final conclusions of the paper, the limitations of the model are explained.

## 2 Methods

### 2.1 Model

We developed a deterministic age-structured compartmental model of SARS-CoV-2 transmission with a population stratified into 5 age groups. The model was constructed by extending the standard SEIR model [3] with additional compartments to model symptomatic and asymptomatic disease progression and to model vaccinated and unvaccinated populations separately.

#### 2.1.1 Extended compartmental SEIR model

We extended the standard SEIR model, by adding compartments to consider different courses of COVID-19 disease, as shown in Figure 1.

**Figure 1:**
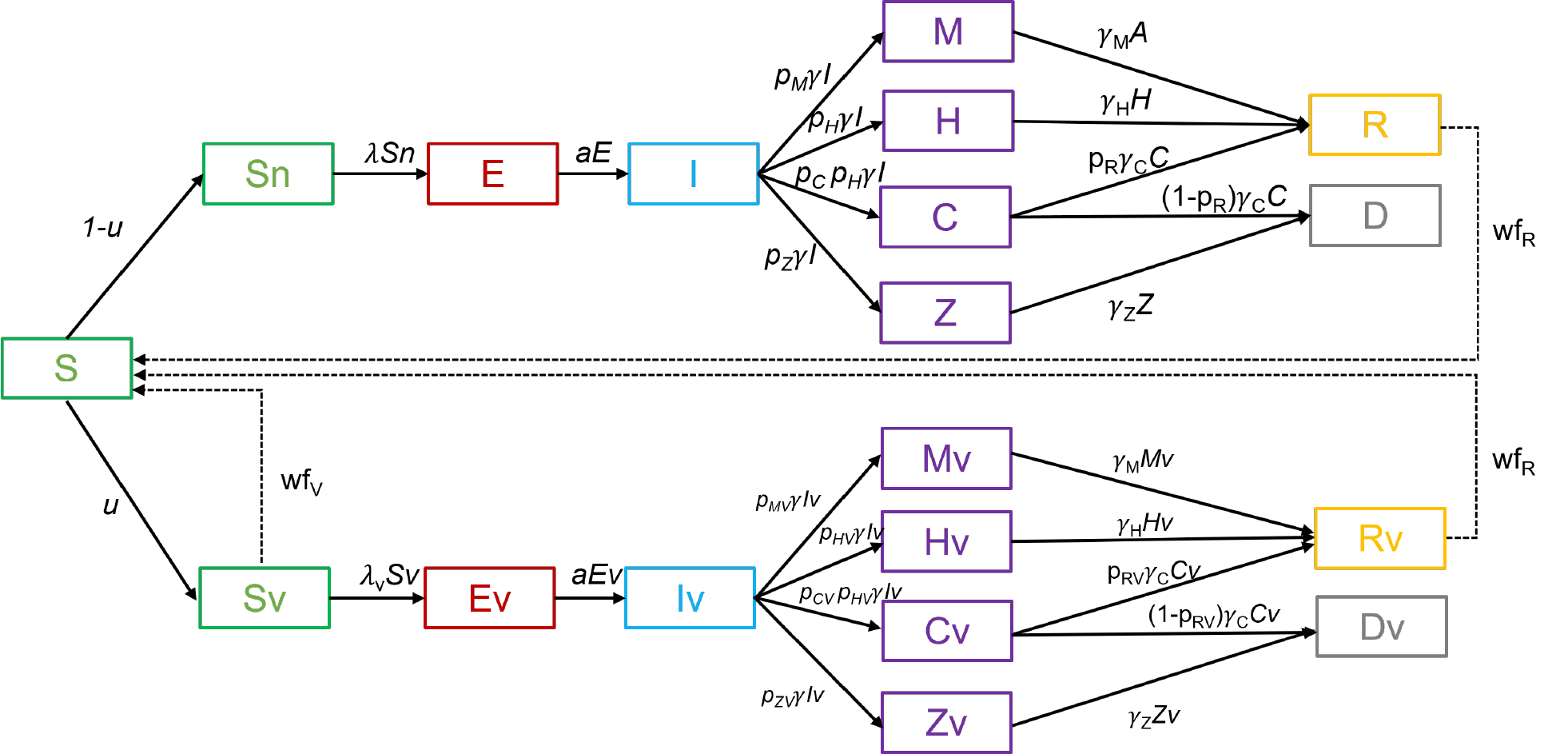
Scheme of the extended SEIR model with additional compartments to model asymptomatic and symptomatic cases with hospital and ICU admissions, split into two separate submodels to model vaccinated and unvaccinated populations.

Those who are susceptible to the disease (compartment S) may become infected and enter the incubation state (compartment E), after some time they become infectious (compartment I) and begin to recover from the infection in different ways. Most people can have an infection with mild symptoms or are asymptomatic (compartment M) and then recover from the infection (compartment R). This is the pathway S→E→I→M→R in the model. Some have severe symptoms requiring hospitalization (compartment H), where they may remain until further recovery (compartment R). This is the pathway S→E→I→H→R in the model. Some of the patients require additional intensive care (compartment C). Patients in the intensive care units may recover and go to compartment R, this is the pathway S→E→I→C→R, or die, this is the pathway S→E→I→C→D in the model. There is an additional compartment Z for modeling people who died from COVID-19 but were not treated in hospitals. This corresponds to the pathway S→E→I→Z→D in the model. This compartment was added during the second wave of COVID-19 in Slovenia in the fall/winter of 2020, when Slovenia witnessed severe outbreaks of SARS-CoV-2 in nursing and retirement homes and could not treat all severely ill people in hospitals. In the following waves of the epidemic, such a course of the disease was rare.

The above idea was applied separately to vaccinated and unvaccinated populations. The joint model was then merged from these two submodels by dividing the susceptible group (S) into two subgroups, Sn and Sv, representing the unvaccinated and vaccinated susceptibles, respectively. The split of the susceptible group into Sn and Sv is defined by the parameter *u*, which represents the proportion of vaccinated at a given time. As can be seen from Figure 1, the two submodels have identical compartments but differ in parameters related to the probability of infection, severity of disease, and possibility of death.

All the parameters of the model are summarized in Table 1. The parameters can be broadly divided into three groups: the proportion parameters, the duration parameters, and the parameters corresponding to the transmission of the infection. The proportion parameters are vaccination rate *u*(*t*), the proportion of mild/asymptomatic infections *p*_*M*_(*t*), the proportion of hospitalizations *p*_*H*_(*t*), the proportion of intensive care hospitalizations *p*_*C*_(*t*), the proportion of hospitalizations resulted in death 1 – *p*_*R*_(*t*), and the proportion of deaths outside hospital care *p*_*Z*_(*t*). All these parameters are estimated daily using data on confirmed positive cases, regular and intensive care hospitalizations, deaths, and vaccination progress. Duration parameters correspond to the latent period duration 1/a(t), the infectious period duration 1/*γ*(*t*), the mild/asymptomatic period duration 1/*γ*(*t*), the intensive care hospitalization 1/*γ*_*M*_(*t*), the non-ICU hospitalization 1/*γ*_*C*_(*t*), and the pre-death duration 1/*γ*_*H*_(*t*) of patients in nursing/retirement homes. This group also includes two durations of waning immunity: the period of waning immunity after disease 1/*wf*_*R*_(*t*) and the period of waning immunity after vaccination 1/*wf*_*V*_(*t*). The time 1/*wf*_*R*_(*t*) corresponds to the mean duration, in which the patient becomes susceptible to re-infection, the time 1/*wf*_*V*_(*t*) represents the mean duration, in which the vaccinated person becomes susceptible to infection. The two durations of waning immunity are set to a longer period, e.g., one year, which means that in one year those who have been vaccinated or recovered from the disease will again become susceptible. Both rates are included in the model in a way to be linearly proportional over time, meaning that after six months, for example, half of those who have been vaccinated or have recovered from the disease will be susceptible again. In this way, we tried to incorporate in the model also the efficacy of the vaccine, which changes over time.

**Table 1:** The parameters of the model. All parameters are functions of time.

| Parameter | Meaning | Value (Range) / Description |
| --- | --- | --- |
| $u(t)$ | proportion of vaccinated | estimated from daily vaccination reports, age-stratified |
| $\lambda(t)$ | force of infection | calculated from transmission coefficients and contact matrices (see age-stratified model) |
| $\frac{1}{a(t)}$ | mean duration of latent period | (3–5) days, depends of SARS-CoV-2 variants |
| $\frac{1}{\gamma(t)}$ | mean infectious period | (3–8) days, depends of SARS-CoV-2 variants |
| $\frac{1}{\gamma_M(t)}$ | mean duration of mild/asymptomatic infection | (7–14) days, depends of SARS-CoV-2 variants |
| $\frac{1}{\gamma_H(t)}$ | mean duration of non-ICU hospitalization | (10–15) days |
|  |  | 14 days |
| $\frac{1}{\gamma_Z(t)}$ | mean duration of the disease before death in nursing/retirement home | |
| $\frac{1}{\gamma_C(t)}$ | mean duration of ICU hospitalization | (14-21) days |
| $p_M(t) = 1 - p_H(t) - p_Z(t)$ | proportion of mild/asymptomatic infections | computed from proportions $p_H(t)$ in $p_Z(t)$, age-stratified |
| $p_H(t)$ | proportion of infections requiring hospitalization | estimated from data, proportions between daily incidence of cases and hospitalizations, age-stratified |
| $p_C(t)$ | proportion of hospitalizations requiring ICU | estimated from hospitalization data reports, proportions between daily incidence of hospitalizations and ICU, age-stratified |
| $p_Z(t)$ | proportion of infections without hospitals resulting in death | estimated for the data in the 2<sup>nd</sup> wave, in all other waves eq. to 0.0, age-stratified |
| $p_R(t)$ | proportion of recovered hospitalizations | estimated from hospitalization data reports, proportions between daily incidence of hospitalizations and deaths, age-stratified |
| $\frac{1}{wf_R(t)}$ | mean duration of waning immunity | (180 - 365) days<br>example: 360 days mean 50% of recovered lost immunity in 180 days. |
| $\lambda_V(t) = \frac{1}{f_{IV}(t)} \cdot \lambda(t)$ | force of infection in the vaccinated group | force of infection in the vaccinated group is equal to force of infection in the unvaccinated group lowered by a certain factor $f_{IV}$<br>$f_{IV}(t)$ is in range (2 - 5),<br>depends of SARS-CoV-2 variants |
| $p_{MV}(t) = 1 - p_{HV}(t) - p_{ZV}(t)$ | proportion of mild/asymptomatic infections in the vaccinated group | computed from proportions $p_{HV}(t)$ in $p_{ZV}(t)$, age-stratified |
| $p_{HV}(t) = \frac{1}{f_{HV}(t)} \cdot p_H(t)$ | proportion of infections requiring hospitalization in the vaccinated group | proportion of hospitalization in the vaccinated group is equal to proportion of hospitalization in the unvaccinated group lowered by a certain factor $f_{HV}$<br>$f_{HV}(t)$ is in range (5 - 10),<br>depends of SARS-CoV-2 variants,<br>age-stratified |
| $p_{CV}(t)$ | proportion of hospitalizations requiring ICU in the vaccinated group | $p_{CV}(t) = p_C(t)$, age-stratified |
| $p_{ZV}(t)$ | proportion of infections without hospitalizations resulting in death in the vaccinated group | 0.0<br>(vaccination starts in Slovenia after the 2nd wave) |
| $1 - p_{RV}(t) = \frac{1}{f_{DV}(t)} \cdot (1 - p_R(t))$ | proportion of hospitalizations resulting in death in the vaccinated group | proportion of hospitalizations resulting in death in the vaccinated group is equal to the proportion of hospitalizations resulting in death in the unvaccinated |
| | | group lowered by the factor $f_{DV}$<br>$f_{DV}(t)$ is in range (5 - 10),<br>depends of SARS-CoV-2 variants,<br>age-stratified |
| $\frac{1}{wf_V(t)}$ | mean duration of waning immunity after vaccination | (180 - 365) days<br>example: 360 days mean 50% of recovered lost immunity in 180 days. |

Disease transmission is included in the model by the parameters *λ*(*t*) corresponding to the force of infection [3], which are computed in the age-stratified extended SEIR model described in the next section.

#### 2.1.2 Age-stratified extended SEIR model

The SEIR model from the previous section has been additionally extended to model five age groups of the population. This was done by cloning a structure of the base model (Figure 1) five times and allowing population mixing across all age subgroups from these submodels. The resulting extended SEIR model is shown in Figure 2.

**Figure 2:**
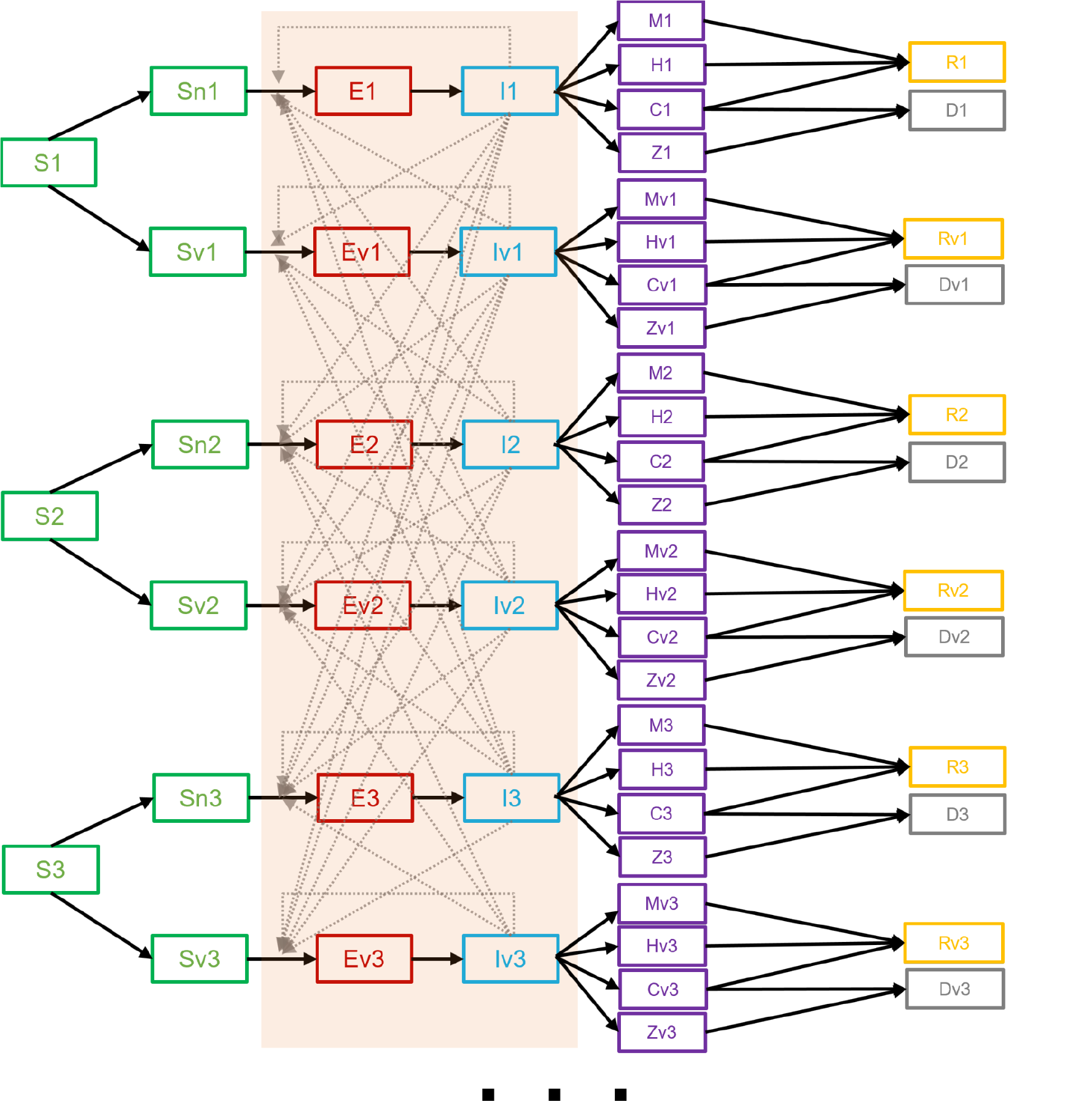
Scheme of the group-stratified extended SEIR model obtained by cloning the base model from Figure 1, which allows for weighted and time-dependent population mixing between groups. The concept was used to model five population age groups.

Each submodel of the model schemed in Figure 2 allows for modeling of the same disease courses and has the same parameters as the base SEIR model shown in Figure 1, whereby the values of the parameters in the submodels may differ. We chose to model five age groups of the population to obtain an age-stratified model that better fits the epidemiological situation with the age-dependent vaccination strategy in Slovenia and the different severity of disease progression within different age groups.

Disease transmission in this model is defined by assuming mixing of populations between age groups and between vaccinated and unvaccinated groups. The force of infection in each submodel is computed as

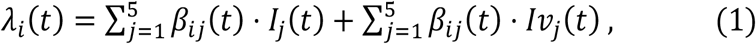

where *λ*_*i*_(*t*) is the force of infection in the submodel representing age group *i*, and is computed as the sum over all age groups of the rates of how many unvaccinated and vaccinated infected individuals from the age group *j*, denoted by *I*_*j*_ and *Iv*_*j*_, infect susceptibles from the age group *i*. The rates for each combination of the groups *i* and *j* are computed as *β*_*ij*_(*t*) ⋅ *I*_*j*_(*t*), where *β*_*ij*_(*t*) represents the contact rate between individuals of group *i* and individuals of group *j*. Contact rates *β*_*ij*_(*t*) are computed as *β*_*ij*_(*t*) = *β*(*t*) ⋅ *w*_*ij*_(*t*), where *β*(*t*) is an overall transmission rate of the disease, and *w*_*ij*_(*t*) are the weights for increasing or decreasing the transmission rate according to the assumed contact mixing of the groups *i* and *j*. The weights are stored in the contact matrix *W*. The overall transmission rate *β*(*t*) is computed as a product of the effective reproduction number *R*(*t*) and the mean infectious period 1/*γ*(*t*), namely *β*(*t*) = *R*(*t*)/*γ*(*t*).

Note that the forces of infection of vaccinated groups, *λν*_*i*_(*t*), are the same as *λ*_*i*_(*t*) reduced by a specific factor *f*_IV_(*t*) that accounts for the effectiveness of vaccination in reducing transmission of the disease (see Table 1).

#### 2.1.3 Model outputs

The model produces the following projections for each age group: 7-day averages of daily positive cases, daily non-intensive care hospitalization admissions and active stays, daily intensive care admissions and active stays, and 7-day averages of deaths. The projections are additionally summarized across all age groups to obtain overall projections. From these projections, we derived cumulative projections of positive cases, hospitalizations, intensive-care hospitalizations and total deaths. In addition, the model also generates weekly incidence of positive cases, hospitalizations and deaths.

#### 2.1.4 Parameters computation and model optimization

The model is calibrated to the current state of the epidemic in Slovenia by adjusting the model parameters in Table 1. All parameters are treated as functions of time, so their values may change over time according to the varying epidemiological situation. While some model parameters are calculated daily, others are estimated by observing the dynamics of the epidemic in the population, and some are set according to the literature and mainly remain fixed over time.

We performed the optimization routine to minimize the error of the model projections on four different objective functions: daily reported infections, daily deaths, daily reported number of current hospitalizations not requiring intensive care and daily reported number of current hospitalizations requiring intensive care. All the data for tunning the parameters were collected from COVID-19 Sledilnik [2] and the Slovenian National Institute of Health [18]. We collated the population into 5 age groups: 0-24, 25-44, 45-64, 65-74, and older than 75. This age grouping was chosen because, from a clinical point of view, these groups represent in some cases the finest granularity we could find in the data, and from a computational point of view, it reduces the dimension of the optimization problem [12].

The parameters estimated daily are the forces of infection *λ*_*i*_(*t*) and the vaccination rates *u*_*i*_(*t*) in each age group *i*. The force of infection *λ*_*i*_(*t*) is calculated according to Eq. (1). For these computations we need the daily estimated effective reproduction number *R*(*t*) weighted by the contact matrix *W*. The reproduction number *R*(*t*) is estimated from the time-series of daily reported infections by using the EpiEstim CRAN R package [19], while the contact matrices *W*(*t*) were set manually by following [20] and corrected over time to follow the current epidemiological interventions in the population. The vaccination rates are computed directly from the data of the daily reported vaccination rates in the collated age groups provided by the Slovenian National Institute of Health [18].

The proportions *p*_*H*_(*t*), *p*_*C*_(*t*), *p*_*Z*_(*t*) and *p*_*R*_(*t*) are also determined daily. The proportion of hospitalizations *p*_*H*_(*t*) is estimated from the ratio of the time-series of daily active confirmed cases and the hospital stays; the proportion of intensive care hospitalizations *p*_*C*_(*t*) is determined from the time-series of intensive care and non-intensive care hospitalizations; and the proportion of hospitalizations resulted in death, 1 – *p*_*R*_(*t*), is computed from the time-series of hospitalization data and reported deaths. The proportion of deaths outside hospital care *p*_*Z*_(*t*) was used in the second wave in Slovenia and was estimated from the time-series data of hospital deaths and all reported deaths due to COVID-19 in Slovenia in that wave. In all other cases it was set to 0. All these proportions are estimated separately for each age group. For example, the proportions of hospitalizations or deaths are much lower in the 0-24 age group than in the 75+ age group. Similarly, the vaccination rates also differ greatly in time between age groups, etc.

The duration parameters 1/a(t), 1/*γ*(*t*), 1/*γ*_*M*_(*t*), 1/*γ*_*C*_(*t*), 1/*γ*_*H*_(*t*), 1/*γ*_*Z*_(*t*) and the durations of waning immunity 1/*wf*_*R*_(*t*), 1/*wf*_*V*_(*t*) are determined manually. They were set according to the literature and remain mainly fixed over time. We changed them only in cases of different SARS-CoV-2 variants when suggested in the literature [21–23].

The optimization and additional calibration of the model are performed daily in the following way. The effective reproduction number *R*(*t*), the proportions *p*_*iH*_(*t*), *p*_*iC*_(*t*), *p*_*iZ*_(*t*), *p*_*iR*_(*t*) and the vaccination rates *u*_*i*_(*t*) are computed daily from the time-series data of daily active confirmed cases, intensive care and non-intensive care hospitalizations, reported deaths and vaccination reports. Several runs of the model are performed, perturbing the parameters around these estimated values (by using perturbation range of ±10%). The resulting model projections are additionally calibrated in each run by automatically adjusting the proportion parameters so that the resulting projections better fit the objective functions on the past data. Typically, 100 runs are made in such an optimization procedure, resulting in 100 instances of model projections. The final projections are created using the median values of these instances. By estimating appropriate quantiles from these instances, we also estimate the 50%- and 95%-confidence intervals of the final projections.

### 2.2 Model evaluation

We have conducted two evaluations of the model: one for the year 2021 (SI-2021 evaluation) and one as part of the European Covid-19 Forecast Hub [17], in which we have participated with our model since December 2021 (EUHub-2022 evaluation).

In both assessments, we focused on three forecast targets: the weekly incidence of new COVID-19 cases, the weekly incidence of new COVID-19 hospitalizations (intensive care and non-intensive care), and the weekly incidence of new COVID-19 deaths in Slovenia. In the SI-2021 evaluation, forecasts were compared with data from COVID-19 Sledilnik [2]. In the EUHub-2022 evaluation, forecasts were compared with data provided by the European Covid-19 Forecast Hub [17], which relies on data from the Johns Hopkins University (for cases and deaths) and data collated by ECDC from national health authorities (for hospitalizations).

Forecasts were made for 1, 2, 3, and 4 weeks ahead, with forecasts calculated each Monday from data up to the last Sunday. Thus, the parameters of the model were estimated from past data, and forecasts were made for up to 4 weeks in advance. Possible future interventions planned by the health authorities to change epidemic dynamics were not considered. Consequently, we assessed the short-term forecasts of the model rather than different possible scenarios according to planned changes in epidemics.

The quality of the forecasts in the SI-2021 evaluation was assessed graphically and by calculating two error statistics. The graphical observations were used to estimate the accuracy of the model over time, while the overall performance of the model was measured by the two error statistics. The absolute differences *RAE*_*i*_ = |*y*_*i*_ − ŷ_*i*_ |/ *y*_*i*_ were calculated for a set of observations *y*_*i*_, *i* = 1, …, *n* and point predictions ŷ_*i*_, *i* = 1, …, *n*. The first error statistic was then computed as the median of *RAE*_*i*_ with interquartile range to compensate for possible outliers in the forecast errors. To obtain a more scale-free error statistic, we compared the model forecasts with baseline forecasts. The baseline forecasts were constructed as forecasts by repeating an observation from the current week four times to predict future observations 4 weeks in advance. The comparison of forecast errors was then measured using a ratio *θ* = (mean of *RAE*_*i*_ of our model) / (mean of *RAE*_*i*_ of baseline model). The ratio *θ* is a measure of the relative performance of our model compared to the baseline model. For *θ* < 1 our model outperforms the baseline model, and for *θ* > 1 the baseline model is better. This measure was developed according to Cramer *et al*. [24].

In the EUHub-2022 evaluation, we reported evaluation results according to the European Covid-19 Forecast Hub reporting style, using the relative weighted interval score (rel.wis) for the evaluation metric, as described in Cramer *et al*. [24]. Comparisons of our model were made with the ensemble and the baseline EuroCOVIDhub models [25], which were evaluated on the Slovenian data.

## 3 Results

### 3.1 Epidemic situation in Slovenia in years 2021 and 2022

Slovenia started 2021 with a peak of COVID-19 cases from a prolonged second wave that had already begun in the summer of 2020, and rapid antigen testing (RAT) and vaccination against COVID-19 were introduced in late 2020 [2,18,26,27].

As shown in Figure 3a, after a brief decline in cases in March 2021, the number of COVID-19 infections, predominantly with the Alpha variant, which was more than 50% more transmissible in humans compared with the original virus, began to rise again. The proportion of vaccinated individuals in Slovenia at that time was approximately 9%, including 5% with completed initial vaccination protocol. To stop the rise in infections, a partial lockdown was imposed in early April, by keeping education and work at a distance, restricting movement, and limiting entry and exit from the country. To visit certain establishments, bars, or shelters, people had to be vaccinated, tested, or show proof of recovery (3G rule). By summer, the number of COVID-19 infections in Slovenia had decreased significantly, after which the measures were relaxed [2,18,26,27].

**Figure 3:**
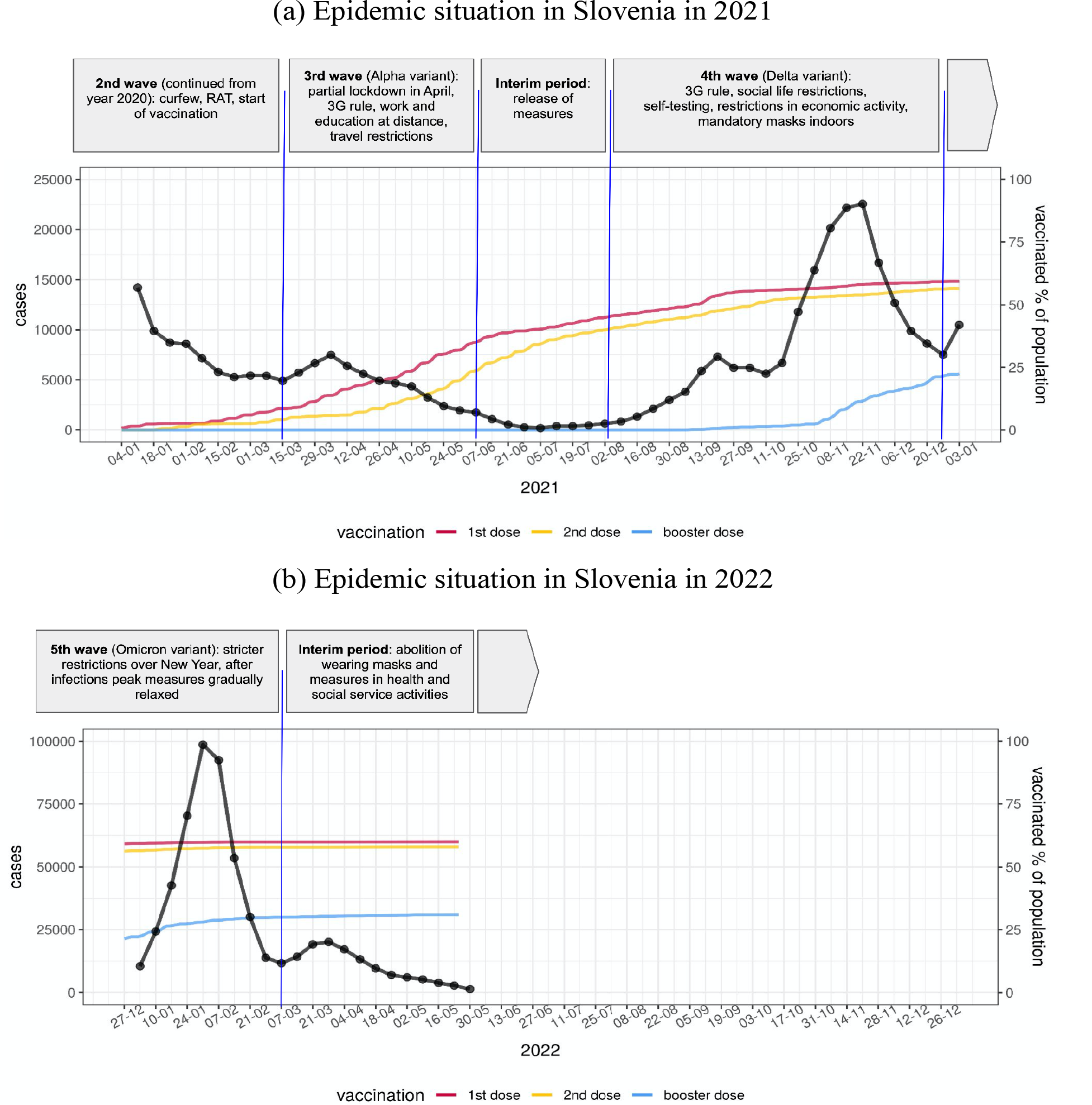
COVID-19 epidemic situation in Slovenia in years 2021 (a) and 2022 (b). Weekly incidence of confirmed COVID-19 cases (black) and the proportion of population vaccinated against COVID-19 with the first (red), the second (yellow), and the booster dose (blue) [2,18,26,27] are shown. The phases of the epidemic, together with prevailing viral variants and non-pharmaceutical public health interventions, are briefly described in the timeline on top.

In August 2021, the number of COVID -19 infections in Slovenia started to rise again. At the beginning of the new school year in September, about 50% of Slovenians were vaccinated, with 45% of the population fully vaccinated. Due to the steady increase in Delta variant infections, the government reintroduced the 3G rule for most of social life on September 15. Measures included restricting social life, limiting the number of people engaged in economic activity, mandatory wearing of surgical or FFP2 masks, and self-testing (3 times per week) for unvaccinated school children, students, and employees in all activities. The increase in infections stalled, but in October the number of new infections began to rise again, reaching a record of more than 22.000 weekly confirmed cases in early November. Slovenia was on the verge of another lockdown as the capacity of 300 beds in intensive care units had almost been reached. The maximum of 288 occupied intensive care unit beds was reached on November 25 [2,18,26,27], without any further implementation of a lockdown. By the end of 2021, 60% of people had been vaccinated, 57% of the population with two doses.

The fifth epidemic wave (Figure 3b) began with the first Omicron variant case confirmed in Slovenia on December 14, and with stricter restrictions over New year holidays. A dramatic increase in infections followed in January 2022, with the wave peaking on January 31 with a weekly incidence of confirmed COVID-19 cases of nearly 100,000 (nearly 5% of the country’s population). It should be noted that despite the record number of confirmed cases, the number of hospitalizations and deaths was not higher than in the previous waves, due in part to the fact that infections with the Omicron variant are often milder than with the Delta variant, and in part to the high vaccination rate in the elderly population. Thereafter, numbers began to decline more rapidly than in previous waves, and measures were gradually relaxed. Entry restrictions and quarantine were lifted, the 3G rule was abolished (except for certain high-risk activities), and a less strict mask-wearing regime was introduced. The percentage of vaccinated individuals had not improved significantly from data at the end of 2021 [2,18,26,27].

### 3.2 Model forecasts evaluation for Slovenia in year 2021

Figures 4, 5, and 6 show model forecasts for the weekly incidence of COVID-19 confirmed cases, hospitalizations, and deaths in Slovenia in 2021, respectively. More detailed versions of these graphs can be found online [28].

**Figure 4:**
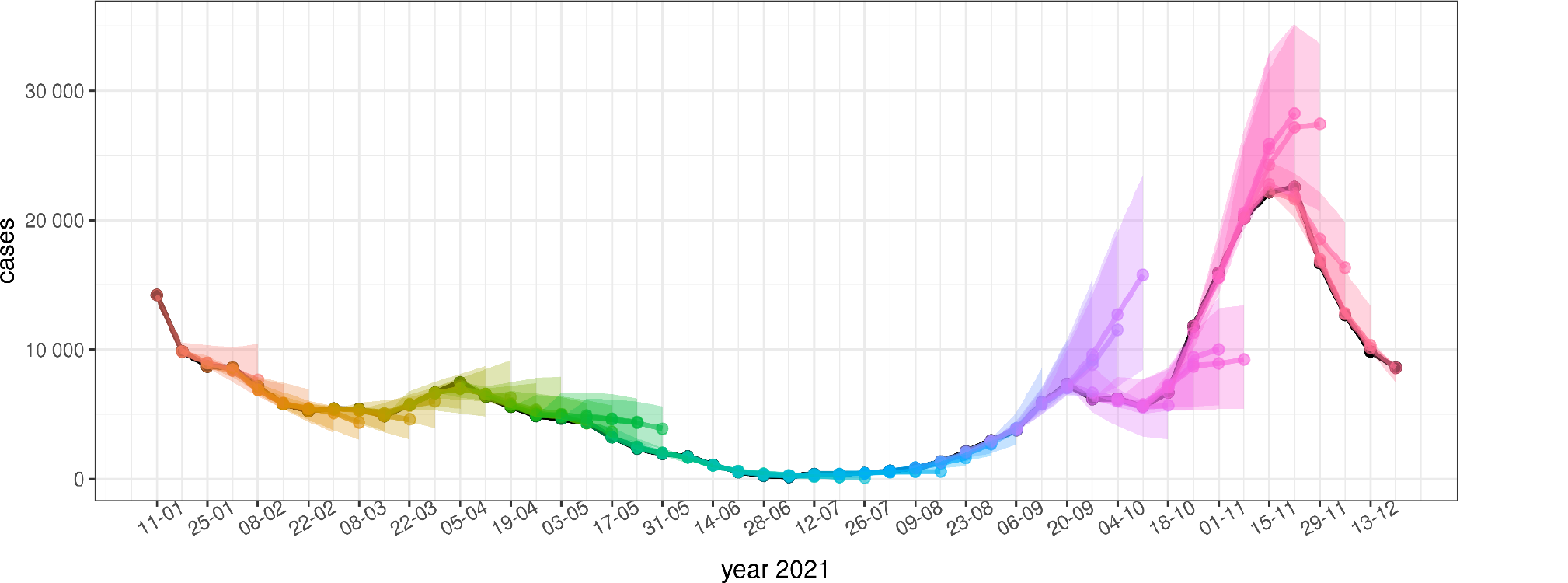
Model forecasts of weekly incidence of COVID-19 confirmed cases. Black dots represent actual data. The colored dots represent predictions for 1, 2, 3 and 4 weeks ahead, the colored areas represent their confidence intervals. Forecasts from the same date are represented with the same color.

**Figure 5:**
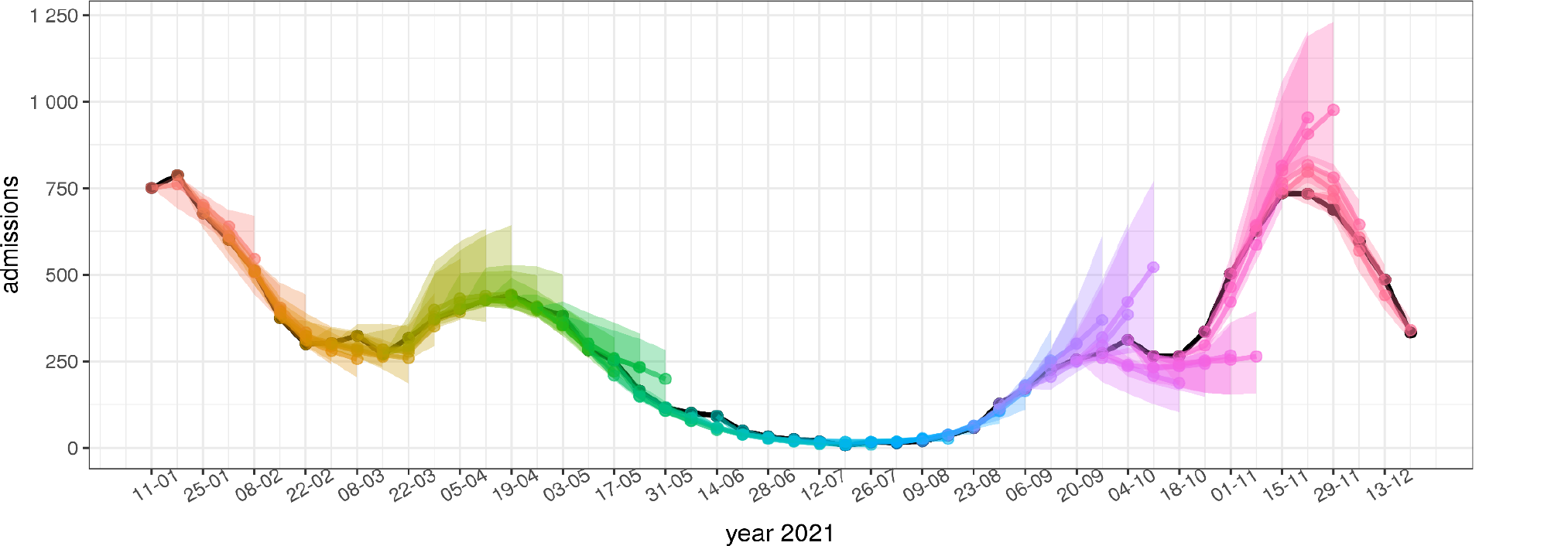
Model forecasts of weekly incidence of COVID-19 hospital admissions. The representation is the same as in Figure 4

**Figure 6:**
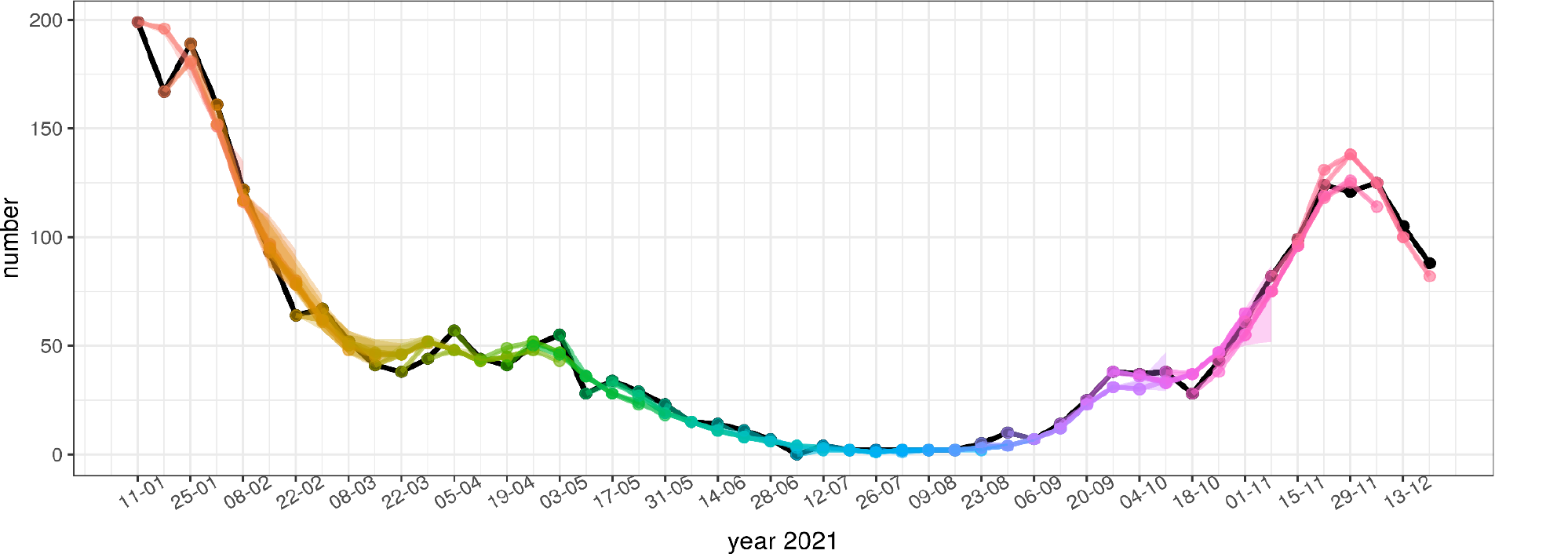
Model forecasts of weekly incidence of deaths due to COVID-19. The representation is the same as in Figure 4.

According to the figures, the accuracy of model forecasts over time depends on the epidemiological situation. The model forecast is less accurate at the beginning and near the peak of the wave, whereas it is more accurate in periods of rising or falling of the wave. This pattern can be clearly seen in the weekly incidence of cases (Fig. 3) and in the weekly incidence of hospital admissions (Fig. 4), while the forecasts of the weekly incidence of deaths (Fig. 6) are more aligned with the actual data even for 4 weeks ahead.

The overall assessment of the model forecasts for the year 2021 is summarized in Table 2. It can be seen that the median of *RAE* for the weekly incidence of confirmed cases, hospitalizations, and deaths is below or around 10% and exceeds this value only for the forecasts of confirmed cases for 4 weeks ahead and the forecasts of hospitalizations for 3 and 4 weeks ahead. The median of *RAE* for the weekly incidence of confirmed cases is only about 3% for the 1-week-ahead forecast, with the Q3 quartile barely reaching 4%. For the longer-term forecasts, the error statistics become larger, as expected, but the median of RAE for the forecast of confirmed cases for 3 weeks ahead is still only about 6%.

**Table 2:** Statistics of the forecast performance of our model in 2021. The first row in the data cells is the median of RAE with interquartile range in square brackets (1st and 3^rd^ quartiles) and the second row is the relative performance of our model compared to the baseline model, measured by the ratio θ.

|  | error type\forecast for | week 1 | week 2 | week 3 | week 4 |
| --- | --- | --- | --- | --- | --- |
| weekly incidence cases | median of $RAE$ [Q1, Q3] | 2.9 [1.6, 4.1] % | 3.2 [1.9, 6.7] % | 6.0 [2.4, 17.8] % | 15.0 [4.8, 51.2] % |
| | Comparison to baseline | $\theta = 0.14$ | $\theta = 0.17$ | $\theta = 0.34$ | $\theta = 0.57$ |
| weekly incidence hospitals | median of $RAE$ [Q1, Q3] | 7.6 [4.1, 12.2] % | 8.5 [4.0, 16.0] % | 11.3 [6.2, 23.2] % | 18.3 [5.9, 35.2] % |
| | Comparison to baseline | $\theta = 0.39$ | $\theta = 0.26$ | $\theta = 0.29$ | $\theta = 0.38$ |
| weekly incidence deaths | median of $RAE$ [Q1, Q3] | 9.8 [3.9, 20.3] % | 9.8 [4.1, 18.3] % | 9.8 [4.2, 18.7] % | 9.8 [4.1, 21.2] % |
| | Comparison to baseline | $\theta = 0.45$ | $\theta = 0.27$ | $\theta = 0.19$ | $\theta = 0.16$ |

The relative performance of the model to the baseline model was measured by the ratio *θ*. The values *θ* are in all forecasts well below 1.0, indicating that our model outperformed the baseline forecasts in all cases. For almost all forecasts in Table 2, the values are even below 0.5, which means that our model almost always provides a forecast that is more than twice as good as the baseline model. For the forecasts of confirmed cases for 1 and 2 weeks ahead and deaths for 3 and 4 weeks ahead, the values *θ* are even below 0.2, which means that we can expect a more than 80% better prediction compared to the baseline model.

### 3.3 Model forecasts evaluation within European Covid-19 Forecast Hub for year 2022

The evaluation of the model for the first three months of 2022 was conducted as part of the European Covid-19 Forecast Hub project. The evaluation results in Table 3 show the relative weighted interval score (rel.wis) for three models: our model (named ULZF-SEIRC19SI in the table), the ensemble model (named EuroCOVIDhub-ensemble in the table) and the baseline model (named EuroCOVIDhub-baseline in the table). The evaluation was performed using data from January 1, 2022 to March 28, 2022. The ensemble is built from all of the models providing forecasts for Slovenia (9 for the weekly incidence of cases, 4 for the weekly incidence of hospital admissions, and 7 for the weekly incidence of deaths). The baseline forecasts are computed in the same way as in the previous evaluation. Note also that the relative interval score is the evaluation measure that compares the performance of the evaluated model to all other models, with values below 1.00 indicating better performance (lower error) of the evaluated model relative to its compared counterparts. More details can be found in Cramer *et al*. [24].

**Table 3:** Performances of the 3 models within European Covid-19 Forecast Hub project on the Slovenian data from January 1, 2022 to March 28, 2022, measured by the relative weighted interval score (rel.wis).

|  | model | rel. wis |  |  |  |
| --- | --- | --- | --- | --- | --- |
|  |  | week1 | week2 | week3 | week4 |
| weekly incidence cases | ULZF-SEIRC19SI | 0.45 | 0.51 | 0.58 | 0.56 |
|  | EuroCOVIDhub-ensemble | 0.58 | 0.62 | 0.72 | 0.77 |
|  | EuroCOVIDhub-baseline | 1.00 | 1.00 | 1.00 | 1.00 |
| weekly incidence hospitals | ULZF-SEIRC19SI | 0.49 | 0.49 | 0.51 | 0.61 |
|  | EuroCOVIDhub-ensemble | 0.61 | 0.55 | 0.48 | 0.64 |
|  | EuroCOVIDhub-baseline | 1.00 | 1.00 | 1.00 | 1.00 |
| weekly incidence deaths | ULZF-SEIRC19SI | 0.73 | 0.58 | 0.47 | 0.56 |
|  | EuroCOVIDhub-ensemble | 0.65 | 0.60 | 0.54 | 0.62 |
|  | EuroCOVIDhub-baseline | 1.00 | 1.00 | 1.00 | 1.00 |

According to the results in Table 3, our model outperformed the ensemble model in almost all forecasts in all three categories. For the forecasts of weekly incidence of cases, our model performed better in all 4 forecasts, with a larger gap in the long-term forecasts. The forecasts of hospital admissions are almost the same for weeks 2, 3, and 4, whereas there is a higher gap in favor of our model for the forecast for 1 week ahead. The weekly incidence of deaths is better forecasted for 1 week ahead with the ensemble model, and for 2, 3, and 4 weeks ahead with our model.

## 4 Discussion

The accuracy of the model over time clearly depends on the epidemiological situation, as can be seen in Figures 2, 3, and 4. The forecasts are less accurate at the beginning and near the peak of the two epidemiologic waves in 2021. Both waves in 2021 were caused by new variants of the SARS-CoV-2 virus with different transmission characteristics. The timing of the introduction of a new viral variant into the population can be speculated, but not accurately predicted. Therefore, forecasts of the onset of waves caused by new variants with different transmission characteristics cannot be accurate. However, when the new viral variant was detected and its transmission properties were incorporated into the model, model forecasts became more reliable and could accurately predict the peak of the wave, assuming no additional public health measures were introduced that could affect epidemiologic dynamics. However, in both waves in 2021 in Slovenia (April 2021 and October-December 2021), a number of public health measures were introduced to slow down the transmission dynamics. Accordingly, at the beginning of the waves, we could not include the changes in transmission due to the measures in the model. As a result, the long-term forecasts predicted peaks without any interventions. The moment the interventions were included in the model, the predictions became more accurate. Errors both at the onset and near the peak of the waves are therefore to be expected and are well known in epidemiological modeling [29–31].

Transmission characteristics of new virus variants were incorporated into the model by estimating the effective reproduction number *R*, whereas interventions were accounted for by changes in the contact matrix *W*. The correct estimation of *R* and contact matrices remains a difficult task and depends strongly on the model structure on the one hand and on the behavior of people during the epidemic on the other [32,33].

It should also be noted that a vaccination campaign was underway in Slovenia in 2021, where 5% of the population, primarily the elderly, was fully vaccinated at the start of the first wave in March 2021, and the second wave started in September 2021 with 45% of the population fully vaccinated. Vaccination was carefully included in the model by dividing the population into age groups and modeling vaccinated and unvaccinated populations separately, following the modeling approach presented by Matrajt *et al*. [11,12]. This allowed us to more accurately include and track the vaccination strategy in Slovenia, resulting in more accurate predictions of hospitalizations and deaths.

Our model also shows an overall better performance compared to the base model and the ensemble model [34] in the European Covid-19 Forecast Hub 2022 evaluation. Although the model is expected to outperform the baseline model, this should not be the case for the ensemble model. Nevertheless, in the model assessment of the first three months of 2022, conducted as part of the European Covid-19 Forecast Hub project, our model performed better than the ensemble model in almost all cases.

The epidemic situation in Slovenia in early 2022 coincided with a large wave of daily confirmed cases that peaked in early February 2022, whereas hospitalizations and deaths did not follow this pattern. This was due to the Omicron variant of the SARS-CoV-2 virus, which is highly transmissible and does not cause as many severe courses of disease as earlier variants [35]. The model could not predict the exact timing of the onset of the Omicron wave, but later, when the model was updated with the features of the new variant and since there were no interventions that caused substantial changes in epidemic dynamics, we were able to accurately predict the timing and height of the peak of the wave as well as the decline. More precise tuning of the proportion parameters in the model and separate vaccination modeling allowed our forecasts to more accurately predict hospitalizations and deaths, although they did not follow the same pattern as in previous waves.

Since our model is one of the models included in the ensemble for Slovenian forecasts within the European Covid-19 Forecast Hub, better results mean that our model forecasts were among the best among all other models. A closer look at the evaluation results of the European Covid-19 Forecast Hub 2022 shows that among the models providing forecasts for Slovenia, our model performed best in all cases of forecasting hospitalizations and deaths, while ranked first for week 1 and second for weeks 2, 3, and 4 for weekly incidence of confirmed cases. The better forecasts in these cases were from the USC-SLKJalpha model [36].

The overall better performance of our model compared with other models for the Slovenian forecasts could be explained by a more appropriate structure of the model for the epidemic situation in Slovenia, in particular by a more refined structure for vaccination and by more and better parameter tuning made possible by the data provided by COVID-19 tracker for Slovenia [2].

## 5 Model Limitations

Our model is a compartmental model, in which the epidemic dynamics within the modeled subgroups of the population are assumed to be homogeneous. We introduced the heterogeneity of transmission to the model by dividing the population into five age groups and modeling the vaccinated and unvaccinated populations separately. But this might not be sufficient to capture the actual dynamics of transmission in the population. This problem could be addressed with other modeling approaches, such as agent-based or network-based modeling [37,38]. However, such modeling introduces a lot of open parameters that need to be estimated using data from different sources (e.g., mobility, localization data, more individual data) that were not available during the epidemics in Slovenia. Therefore, we opted for a less complex and well-established compartmental model, but with extensions that allowed us to increase the model’s accuracy by adding the compartments and parameters that can be estimated from the data we have.

Nevertheless, our model has more than 100 parameters that need to be properly determined. Since many parameters in the model are arbitrarily estimated and may not fully reflect the actual epidemic situation, we attempted to compensate for this by performing an additional calibration of the proportion parameters in the model optimization process. This calibration tries to change the proportion parameters of the model (e.g., the proportion of hospitalizations, the proportion of intensive care, deaths, etc.) to better fit the model projections to the actual data, even if the parameters would no longer reflect the actual situation in the data. In this way, we achieve a better fit of the model projections to the current and past data and compensate for some loosely/arbitrarily estimated open parameters of the model.

In addition, the model forecasts rely only on current data, and we did not attempt to incorporate any expected future interventions into the model, even if they could be predicted based on the model projections. Therefore, the forecasts tend to predict what would happen without possible future interventions or other unexpected situations. In daily operational runs of the model, we also created additional scenarios for situations in which a new epidemiologic wave or a new variant of the SARS-CoV-2 virus was expected. Note that epidemiological forecasts and other epidemiological statistics for Slovenia are calculated daily with our model starting in April 2020. All the results can be found on our web page [7].

## 6 Conclusion

The presented compartmental model is used for modeling the COVID-19 epidemic in Slovenia. The SEIR model was extended by dividing the population into five age groups and allowing separate modeling of vaccinated and unvaccinated populations, to better account for the vaccination strategy in Slovenia as well as various courses of the disease and transmissibility caused by different SARS-CoV-2 variants. The model was extended to the complexity at which the model parameters can still be reliably estimated from the epidemiological data available in Slovenia.

Despite the known limitations of such modeling, we were able to obtain acceptable forecast results for short-term forecasts for up to 4 weeks in advance. Evaluation of model forecasts for 2021 showed the expected behavior of epidemiological modeling: if we do not interfere with disease dynamics, the model predicts the situation well; the changes in epidemiologic dynamics due to the emergence of a new viral variant in the population or the introduction of new interventions cannot be predicted by the model, but when the new situation is incorporated into the model, the forecasts are again reliable.

Comparison of model forecasts with the ensemble forecasts for 2022 within the European Covid-19 Forecast Hub showed better performance of our model, which can be explained by a more appropriate structure of the model for the epidemic situation in Slovenia, in particular a more refined structure for vaccination, and better parameter tuning enabled by the more comprehensive data for Slovenia included in our modeling.

The model has been fully operational on a daily basis since April 2020 and served as one of the models for decision-making during the COVID-19 epidemic in Slovenia. The model is also part of the European Covid-19 Forecast Hub, coordinated by the European Centre for Disease Prevention and Control.

## Data Availability

All data produced are available online at https://covid-19.sledilnik.org/ and https://github.com/sledilnik. Source code of the model described in this study can be found at https://github.com/janezz25/SEIR-C19-SI .

https://apps.lusy.fri.uni-lj.si

## Availability of data and material

The data that support the findings of this study are openly available at https://covid-19.sledilnik.org/ and https://github.com/sledilnik. Sources of the model described in this study can be found at https://github.com/janezz25/SEIR-C19-SI.

## References

1. World Health Organization. Coronavirus disease (COVID-19) pandemic. [Internet]. 2022 [cited 2022 Mar 26]. Available from: https://www.who.int/emergencies/diseases/novel-coronavirus-2019/

2. COVID-19-sledilnik. [Internet]. 2020 [cited 2022 Mar 26]. Available from: https://covid-19.sledilnik.org/

3. Keeling MJ, Rohani P. Modeling Infectious Diseases in Humans and Animals [Internet]. Princeton University Press; 2008 [cited 2022 Mar 26]. Available from: https://www.degruyter.com/document/doi/10.1515/9781400841035/html

4. Leskovar M, Cizelj L. Robust and Intuitive Model for COVID-19 Epidemic in Slovenia. Strojniški vestnik - Journal of Mechanical Engineering. 2022;68:213–24.

5. Zaplotnik Ž, Gavrić A, Medic L. Simulation of the COVID-19 epidemic on the social network of Slovenia: Estimating the intrinsic forecast uncertainty. Shaman J, editor. PLOS ONE. 2020;15:e0238090.

6. Manevski D, Ružić Gorenjec N, Kejžar N, Blagus R. Modeling COVID-19 pandemic using Bayesian analysis with application to Slovene data. Math Biosci. 2020;329:108466.

7. Žibert J. COVID-19 SI [Internet]. APPS LUSY. 2022 [cited 2022 May 15]. Available from: https://apps.lusy.fri.uni-lj.si/

8. Barbarossa MV, Fuhrmann J, Meinke JH, Krieg S, Varma HV, Castelletti N, et al. The impact of current and future control measures on the spread of COVID-19 in Germany [Internet]. Epidemiology; 2020 [cited 2022 Jun 16]. Available from: http://medrxiv.org/lookup/doi/10.1101/2020.04.18.20069955

9. Li ML, Bouardi HT, Lami OS, Trikalinos TA, Trichakis NK, Bertsimas D. Forecasting COVID-19 and Analyzing the Effect of Government Interventions [Internet]. Epidemiology; 2020 [cited 2022 Jun 16]. Available from: http://medrxiv.org/lookup/doi/10.1101/2020.06.23.20138693

10. Khailaie S, Mitra T, Bandyopadhyay A, Schips M, Mascheroni P, Vanella P, et al. Development of the reproduction number from coronavirus SARS-CoV-2 case data in Germany and implications for political measures. BMC Med. 2021;19:32.

11. Matrajt L, Eaton J, Leung T, Brown ER. Vaccine optimization for COVID-19: Who to vaccinate first? Sci Adv. 2021;7:eabf1374.

12. Matrajt L, Eaton J, Leung T, Dimitrov D, Schiffer JT, Swan DA, et al. Optimizing vaccine allocation for COVID-19 vaccines shows the potential role of single-dose vaccination. Nat Commun. 2021;12:3449.

13. Parolini N, Dede’ L, Ardenghi G, Quarteroni A. Modelling the COVID-19 epidemic and the vaccination campaign in Italy by the SUIHTER model. Infect Dis Model. 2022;7:45–63.

14. Chowdhury R, Heng K, Shawon MSR, Goh G, Okonofua D, Ochoa-Rosales C, et al. Dynamic interventions to control COVID-19 pandemic: a multivariate prediction modelling study comparing 16 worldwide countries. Eur J Epidemiol. 2020;35:389–99.

15. Grimm V, Mengel F, Schmidt M. Extensions of the SEIR model for the analysis of tailored social distancing and tracing approaches to cope with COVID-19. Sci Rep. 2021;11:4214.

16. Sjödin H, Johansson AF, Brännström Å, Farooq Z, Kriit HK, Wilder-Smith A, et al. COVID-19 healthcare demand and mortality in Sweden in response to non-pharmaceutical mitigation and suppression scenarios. Int J Epidemiol. 2020;49:1443–53.

17. European Covid-19 Forecast Hub [Internet]. [cited 2021 Dec 12]. Available from: https://covid19forecasthub.eu

18. National Institute of Public Health Slovenia. [Internet]. 2022 [cited 2022 Dec 12]. Available from: https://www.nijz.si/sl/dnevno-spremljanje-okuzb-s-sars-cov-2-covid-19

19. Cori A, Ferguson NM, Fraser C, Cauchemez S. A new framework and software to estimate time-varying reproduction numbers during epidemics. Am J Epidemiol. 2013;178:1505–12.

20. Prem K, Cook AR, Jit M. Projecting social contact matrices in 152 countries using contact surveys and demographic data. Halloran B, editor. PLOS Comput Biol. 2017;13:e1005697.

21. Lauer SA, Grantz KH, Bi Q, Jones FK, Zheng Q, Meredith HR, et al. The Incubation Period of Coronavirus Disease 2019 (COVID-19) From Publicly Reported Confirmed Cases: Estimation and Application. Ann Intern Med. 2020;172:577–82.

22. European Centre for Disease Prevention and Control. Updated projections of COVID-19 in the EU/EEA and the UK. [Internet]. 2020 Nov. Available from: https://www.ecdc.europa.eu/sites/default/files/documents/covid-forecasts-modelling-november-2020.pdf

23. Zhang J, Litvinova M, Liang Y, Wang Y, Wang W, Zhao S, et al. Changes in contact patterns shape the dynamics of the COVID-19 outbreak in China. Science. 2020;368:1481–6.

24. Cramer EY, Ray EL, Lopez VK, Bracher J, Brennen A, Castro Rivadeneira AJ, et al. Evaluation of individual and ensemble probabilistic forecasts of COVID-19 mortality in the United States. Proc Natl Acad Sci U S A. 2022;119:e2113561119.

25. European Covid-19 Forecast Hub: Community [Internet]. 2022 [cited 2022 Jun 6]. Available from: https://covid19forecasthub.eu/community.html

26. Portal GOV.SI [Internet]. 2022 [cited 2022 May 8]. Available from: https://www.gov.si/

27. Our World in Data: Slovenia [Internet]. 2022 [cited 2022 May 15]. Available from: https://ourworldindata.org/coronavirus/country/slovenia.

28. Žibert J. Model evaluation [Internet]. APPS LUSY. 2022 [cited 2022 Jun 16]. Available from: https://apps.lusy.fri.uni-lj.si/~janezz/analize/eval_model_2021.html

29. Roberts M, Andreasen V, Lloyd A, Pellis L. Nine challenges for deterministic epidemic models. Epidemics. 2015;10:49–53.

30. Castro M, Ares S, Cuesta JA, Manrubia S. The turning point and end of an expanding epidemic cannot be precisely forecast. Proc Natl Acad Sci U S A. 2020;117:26190–6.

31. Crépey P, Noël H, Alizon S. Challenges for mathematical epidemiological modelling. Anaesth Crit Care Pain Med. 2022;41:101053.

32. Melegaro A, Jit M, Gay N, Zagheni E, Edmunds WJ. What types of contacts are important for the spread of infections?: using contact survey data to explore European mixing patterns. Epidemics. 2011;3:143–51.

33. Fumanelli L, Ajelli M, Manfredi P, Vespignani A, Merler S. Inferring the structure of social contacts from demographic data in the analysis of infectious diseases spread. PLoS Comput Biol. 2012;8:e1002673.

34. Sherratt K, Bosse N, Funk S. EuroCOVIDhub-ensemble [Internet]. Available from: https://raw.githubusercontent.com/epiforecasts/covid19-forecast-hub-europe/main/data-processed/EuroCOVIDhub-ensemble/metadata-EuroCOVIDhub-ensemble.txt

35. European Centre for Disease Prevention and Control. Assessment of the further spread and potential impact of the SARS-CoV-2 Omicron variant of concern in the EU/EEA, 19th update [Internet]. 2022 [cited 2022 May 15]. Available from: https://www.ecdc.europa.eu/en/publications-data/covid-19-omicron-risk-assessment-further-emergence-and-potential-impact.

36. Srivastava A, Xu T, Prasanna VK. Fast and Accurate Forecasting of COVID-19 Deaths Using the SIkJα Model. [Internet]. arXiv; 2020 [cited 2022 May 15]. Available from: http://arxiv.org/abs/2007.05180

37. Perez L, Dragicevic S. An agent-based approach for modeling dynamics of contagious disease spread. Int J Health Geogr. 2009;8:50.

38. Firth JA, Hellewell J, Klepac P, Kissler S, CMMID COVID-19 Working Group, Kucharski AJ, et al. Using a real-world network to model localized COVID-19 control strategies. Nat Med. 2020;26:1616–22.

